# Protocol: Identification and evaluation of critical factors in achieving high and sustained childhood immunization coverage in selected low- and lower-middle income countries

**DOI:** 10.1101/2021.12.01.21267018

**Authors:** Robert A. Bednarczyk, Kyra A. Hester, Sameer M. Dixit, Anna S. Ellis, Cam Escoffery, William Kilembe, Katie Micek, Zoë Sakas, Moussa Sarr, Matthew C. Freeman, the Vaccine Exemplars Research Consortium

**Author notes:** Corresponding author: Name: Robert A Bednarczyk, Mailing address: Rollins School of Public Health, Emory University, 1518 Clifton Road NE, Atlanta, GA 30322, USA. Vaccine Exemplars Research Consortium Co-Authors: Natália S. Bueno, Bonheur Dounebaine, Kimberley R. Isett, Pinar Keskinocak, B. Pablo Montagnes, Dima Nazzal, Saad Omer, Walter Orenstein, Miguel R. Robayo, Simone Rosenblum, Francisco Castillo Zunino Note that all authors have seen and approved of this manuscript.

## Abstract

**Introduction:** Increases in global childhood vaccine delivery have led to decreases in morbidity from vaccine-preventable diseases. However, these improvements in vaccination have been heterogeneous, with some countries demonstrating greater levels of change and sustainability. Understanding what these high-performing countries have done differently and how their decision-making processes will support targeted improvements in childhood vaccine delivery.

**Methods and analysis:** We studied three countries - Nepal, Senegal, Zambia - with exemplary improvements in coverage between 2000-2018 as part of the Exemplars in Global Health Program. We apply established implementation science frameworks to understand the “how” and “why” underlying improvements in vaccine delivery and coverage. Through mixed methods research we will identify drivers of catalytic change in vaccine coverage and the decision-making process supporting these interventions and activities. Methods include quantitative analysis of available datasets and in-depth interviews and focus groups with key stakeholders in the global, national, and sub-national government and non-governmental organization space, as well as community members and local health delivery system personnel.

**Ethics and dissemination:** Working as a multinational and multidisciplinary team, and under oversight from all partner and national-level (where applicable) institutional review boards, we collect data from participants who provided informed consent. Findings are disseminated through a variety of forms, including peer-reviewed manuscripts related to country-specific case studies and vaccine system domain-specific analyses, presentations to key stakeholders in the global vaccine delivery space, and narrative dissemination on the Exemplars.Health website.

**Strengths and limitations of this study:**

- This study is led by a multidisciplinary team and grounded in several theoretical frameworks across disciplines from implementation science to behavioral theory.
- We utilized a cross cutting, cross-disciplinary, approach, which assessed relevant domains across our selected exemplars countries as well as within the subjects that arise from the data, over a roughly 20-year time horizon.
- We selected three countries with historically high unvaccinated populations to represent different geographies, cultures, and governments, as well as to highlight regions with historically high unvaccinated populations.
- We did not study a less successful, or “non-exemplar”, counterfactual country.
- The research tools identified and explored catalytic events and the implementation of external policies and development of internal policies and systems, with a focus on participants’ current experiences and perceptions of prior activities.

## Introduction

Early childhood vaccination is widely recognized as one of the most important public health interventions. Increasing vaccine coverage globally has substantially reduced the incidence of, and mortality from, vaccine-preventable diseases.^[1]^ While early childhood vaccine coverage has increased globally, there are still millions of children, particularly in low- and lower-middle-income countries (LICs and LMICs, respectively), who remain unvaccinated.^[2]^ The World Health Organization’s (WHO) Global Vaccine Action Plan (GVAP) sets global targets for all countries to achieve 90% national level coverage of diphtheria, tetanus, pertussis (DTP) for three doses of vaccine (DTP3), and 80% sub-national level DTP3 coverage in every district by 2015.^[3, 4]^ Although significant progress has been made toward these goals – global DTP3 coverage increased from 72% in 2000 to 86% in 2018 – the WHO/UNICEF Estimates of National Immunization Coverage (WUENIC) demonstrate that this progress fell short in both coverage and equity.^[5]^ The COVID-19 pandemic has also negatively impacted routine immunization globally; the extent of this impact is still being assessed,^[6-8]^ and is outside of the scope of this retrospective evaluation.

The literature documenting identified barriers and facilitators of improved vaccine coverage is vast. The systematic review performed by Phillips et al. (2017) provides a conceptual framework identifying facility readiness, intent to vaccinate, and community access as the core determinants of effective vaccine coverage.^[9]^ Similarly, LaFond et al. (2015) identified direct and enabling drivers of immunization coverage improvement as well as essential health and immunization system components, such as district management teams and existence of basic routine immunization resources and capacity.^[10]^

Identification of these barriers and facilitators is only a first step towards improving global vaccine coverage. There remains an evidence gap in understanding “how” and “why” these factors influence system performance. Notably, to strengthen immunization program function we need to understand the development, implementation, and adaptation of programs and interventions. Little rigorous evidence is available on the specific paths to success, including implementation strategies, in the LICs and LMICs that have achieved high and sustained immunization coverage.

We apply a “positive deviant” approach to study high-performing countries, i.e., to understand successful vaccine system performance by identifying positive outliers – countries or systems that exceed their peers – and studying the factors that supported catalytic growth to reach a high level of coverage.^[11]^ Through the identification of the components and pathways to high vaccine coverage among exemplar countries, actionable recommendations can be developed and disseminated to other countries that have not yet had similar success. These recommendations can support decision-making processes to improve immunization programs and health systems, improve overall vaccine coverage, and mitigate inequities in sub-national vaccine coverage in these countries.

The Exemplars in Vaccine Delivery - nested within the larger Exemplars in Global Health partnership, aims to identify the “how” and “why” behind implementation of particular systems and decisions that led to high and sustained infant vaccine coverage through a geographically diverse set of positive deviant countries (i.e., Nepal, Senegal, Zambia).^[12]^ Using two complementary implementation science frameworks and a multi-disciplinary approach - reaching beyond medical and public health research - we built on the existing evidence and frameworks to explore specific components or critical factors of the immunization system to identify potential areas of future research and investment in immunization system interventions. This manuscript presents our mixed methods data collection methods to address these outstanding questions.

## Methods and analysis

### Overview

The purpose of this study is to assess “how” and “why” some countries have succeeded in achieving significantly improved coverage rates between 2000-2018, and to provide actionable recommendations for improving national and sub-national vaccine coverage. This study focuses on three areas of inquiry: (1) Critical policy and programmatic innovations that drove changes to vaccine coverage and equity; (2) “how” and “why*”* these innovations were implemented; and (3) cross-country syntheses of key success factors.

Our research consortium includes Emory University, Georgia Institute of Technology, the University of Delaware, the Center for Molecular Dynamics in Nepal (CMDN), the Center for Family Health Research in Zambia (CFHRZ), the Institut de Recherche en Santé de Surveillance Epidemiologique et de Formation (IRESSEF; Institute for Health Research, Epidemiological Surveillance, and Training) in Senegal.

### Selection of Exemplar Countries

Three exemplar countries – Nepal, Zambia, and Senegal - were selected based on available data and expert review:. Countries were eligible for inclusion if, in the year 2000, (a) their population exceeded 5 million, and (b) the World Bank classified them as low income. Forty-seven countries met these criteria. Two analyses were performed to identify exemplars from the eligible countries based on measured coverage of DTP1 and DTP3: direct estimates of the compound annual growth rate (CAGR) of vaccine coverage over 5-year increments and a segmentation analysis based on coverage, dropout rates, and country conflict status (Figure 1). Taken together, DTP1 and DTP3 serve as common proxies for the function of the vaccine delivery system in each country, as DTP1 can indicate how many children are reached by the immunization system, and DTP3 can indicate how many children the program has continued to reach.

**Figure 1.**
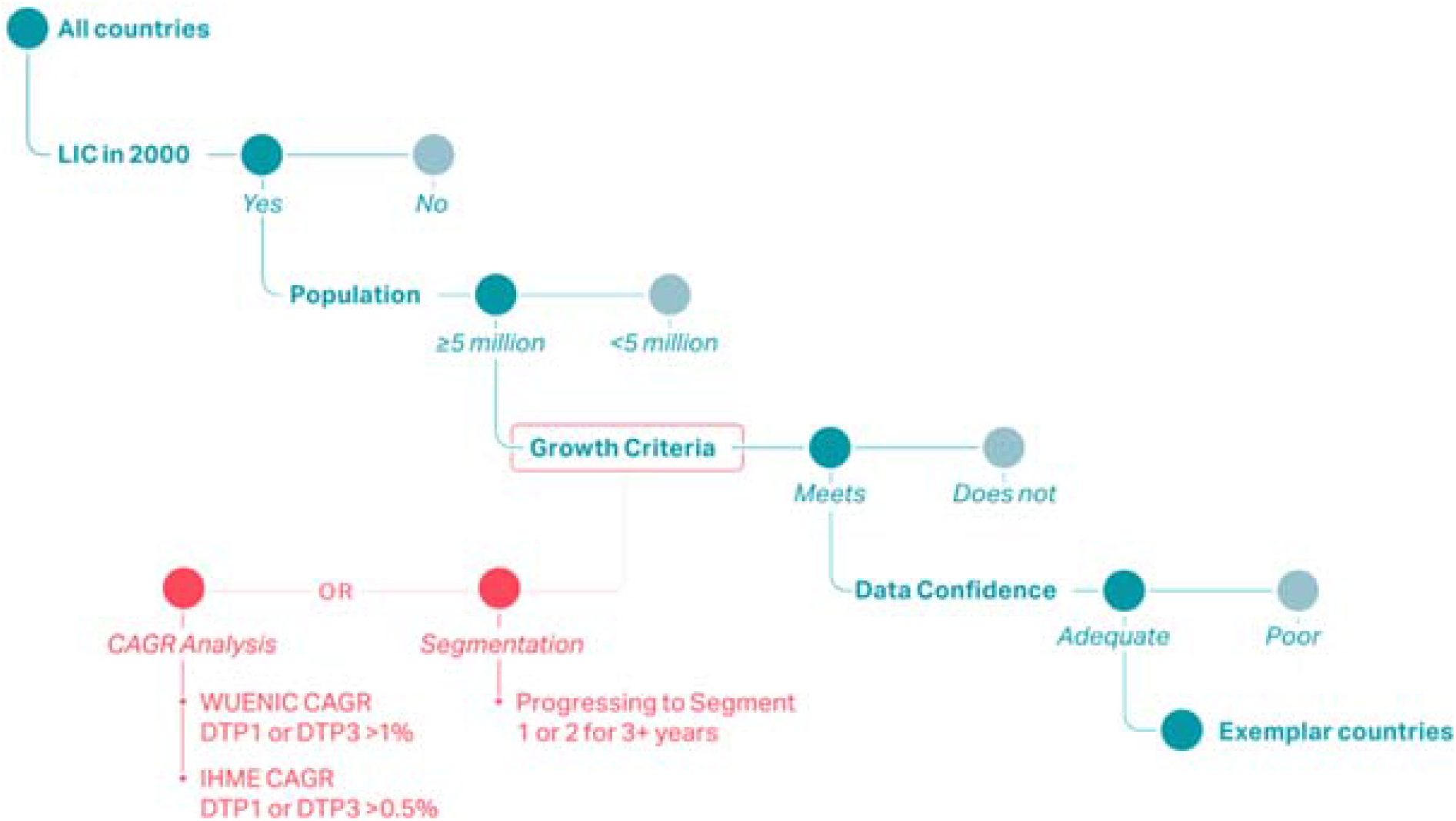
Country filtering process, of which 47 countries met the growth criteria.

The CAGR analysis utilized both WUNEIC and Institute of Health Metrics and Evaluation (IHME) data.^[5, 13]^ For the above mentioned 47 countries, we calculated CAGRs for each country, with both WUENIC and IHME data, from 2000-2016. CAGR calculations used three-year rolling averages. We found the highest-performing countries by applying pre-determined cutoffs by data source; the cutoff percentage depended on the overall performance of the group. The WUENIC data had a CAGR cutoff of 0.9%, indicating a 9% increase over 10 years, and the IHME data had a CAGR cutoff of 0.5%, indicating a 5% increase over 10 years. Seventeen countries met both the WUENIC and IHME CAGR cutoff percentage.

The segmentation analysis used the rolling three-year averages obtained from WUENIC data. ^10^ Five segments were created by analyzing and ranking DTP1 coverage, DTP3 coverage, dropout rates, and conflict. The segments were classified as follows: **Segment 1** countries had ‘proven themselves’ with national DTP3 coverage greater than 90%; **Segment 2** included countries that were ‘on the right track’ with national coverages of DTP3 less than or equal to 90%, but DTP1 greater than 80% and a dropout rate greater than 10%; **Segment 3** included countries that were ‘getting children back into the system,’ with national coverages of DTP3 90%, DTP1 80%, and a dropout rate 10%; **Segment 4** included countries that were still ‘building essentials’, with national coverages of DTP3 90%, DTP1 80%, and no conflict at time of selection; and **Segment 5** included countries with ongoing conflict at time of selection. Exemplar countries were identified as those meeting all three of the following criteria: (1) The country was in segment 3, 4, or 5 at any time during the period 2005-2010; (2) The country progressed to either segment 1 or 2; and (3) The country stayed in segment 1 or 2 for at least 3 years (Figure 2).

**Figure 2.**
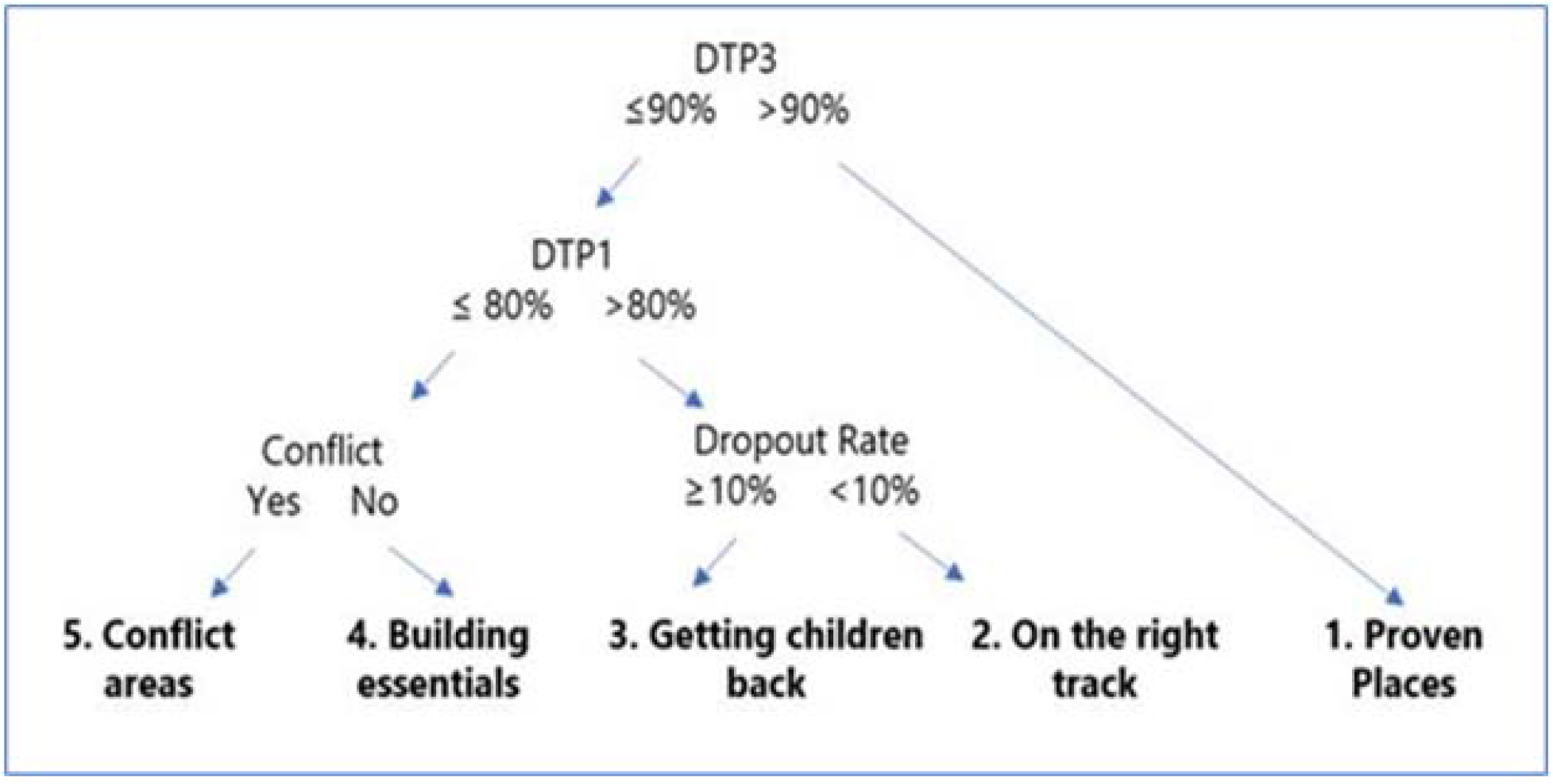
Segment analysis logic.

The shortlist of possible exemplar countries, based on both analyses, had 13 countries (Table 1). The final three countries were selected to represent geographic diversity (South Asia, East Africa, West Africa), as these regions have the majority of unvaccinated children globally. The democracy index, as defined by the 2018 Democracy Index, was used for framing the country selection, and for exclusion criteria.^[14]^

**Table 1.**
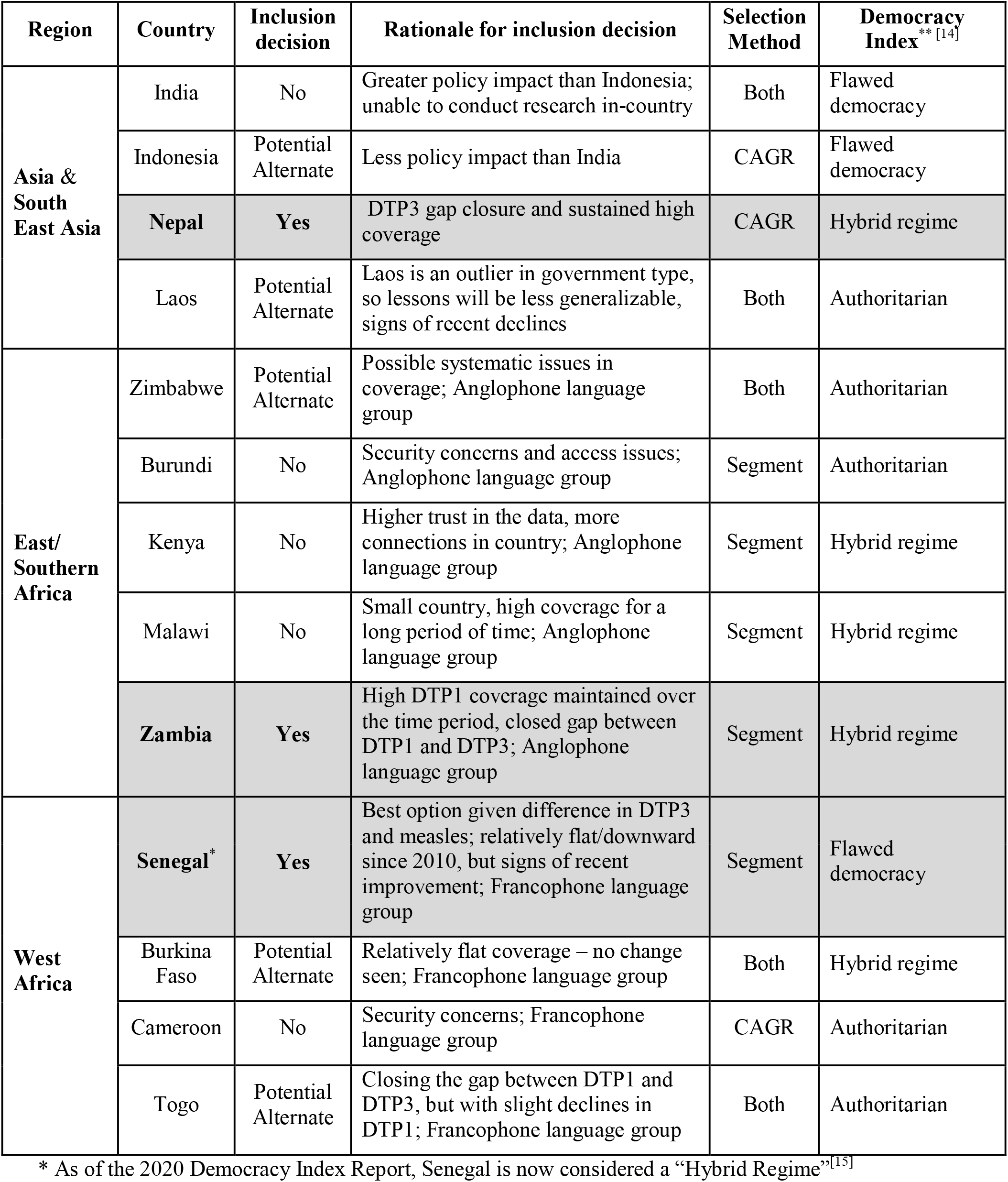
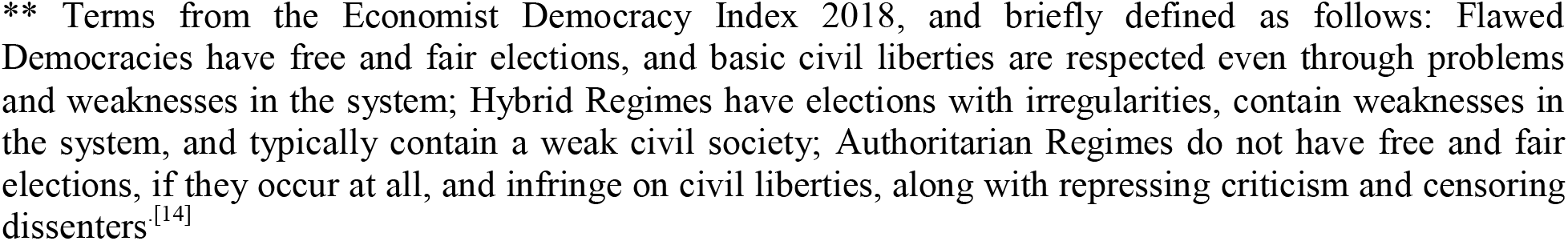
Additional country selection criteria considered during study planning, and rationale for final selection, as of 2018

| Region | Country | Inclusion decision | Rationale for inclusion decision | Selection Method | Democracy Index<sup>**</sup> [14] |
| --- | --- | --- | --- | --- | --- |
| Asia & South East Asia | India | No | Greater policy impact than Indonesia; unable to conduct research in-country | Both | Flawed democracy |
|  | Indonesia | Potential Alternate | Less policy impact than India | CAGR | Flawed democracy |
|  | **Nepal** | **Yes** | DTP3 gap closure and sustained high coverage | CAGR | Hybrid regime |
|  | Laos | Potential Alternate | Laos is an outlier in government type, so lessons will be less generalizable, signs of recent declines | Both | Authoritarian |
| East/ Southern Africa | Zimbabwe | Potential Alternate | Possible systematic issues in coverage; Anglophone language group | Both | Authoritarian |
|  | Burundi | No | Security concerns and access issues; Anglophone language group | Segment | Authoritarian |
|  | Kenya | No | Higher trust in the data, more connections in country; Anglophone language group | Segment | Hybrid regime |
|  | Malawi | No | Small country, high coverage for a long period of time; Anglophone language group | Segment | Hybrid regime |
|  | **Zambia** | **Yes** | High DTP1 coverage maintained over the time period, closed gap between DTP1 and DTP3; Anglophone language group | Segment | Hybrid regime |
| West Africa | **Senegal\*** | **Yes** | Best option given difference in DTP3 and measles; relatively flat/downward since 2010, but signs of recent improvement; Francophone language group | Segment | Flawed democracy |
|  | Burkina Faso | Potential Alternate | Relatively flat coverage – no change seen; Francophone language group | Both | Hybrid regime |
|  | Cameroon | No | Security concerns; Francophone language group | CAGR | Authoritarian |
|  | Togo | Potential Alternate | Closing the gap between DTP1 and DTP3, but with slight declines in DTP1; Francophone language group | Both | Authoritarian |

\* As of the 2020 Democracy Index Report, Senegal is now considered a “Hybrid Regime”<sup>[15]</sup>
\*\* Terms from the Economist Democracy Index 2018, and briefly defined as follows: Flawed Democracies have free and fair elections, and basic civil liberties are respected even through problems and weaknesses in the system; Hybrid Regimes have elections with irregularities, contain weaknesses in the system, and typically contain a weak civil society; Authoritarian Regimes do not have free and fair elections, if they occur at all, and infringe on civil liberties, along with repressing criticism and censoring dissenters.<sup>[14]</sup>

### Country-level data collection

We conducted research at different levels of the healthcare system for each country: the national level, three sub-national regions/provinces, and three districts per region/province for a total of nine districts. Our pre-determined sub-national region selection criteria differed by country, but one region in each country contained the capital city of the country, with the other two regions stratified on factors determined with input from the local study team (e.g., high/low sub-national immunization coverage, rural/urban, road access/lack of road access, ethnic/religious minority/majority). Changes in sub-national immunization coverage over time were assessed using district-level data (Figures 3A, 3B, 3C). Districts were selected based on country specific CAGR and DTP3 percentile cutoffs.

**Figure 3.**
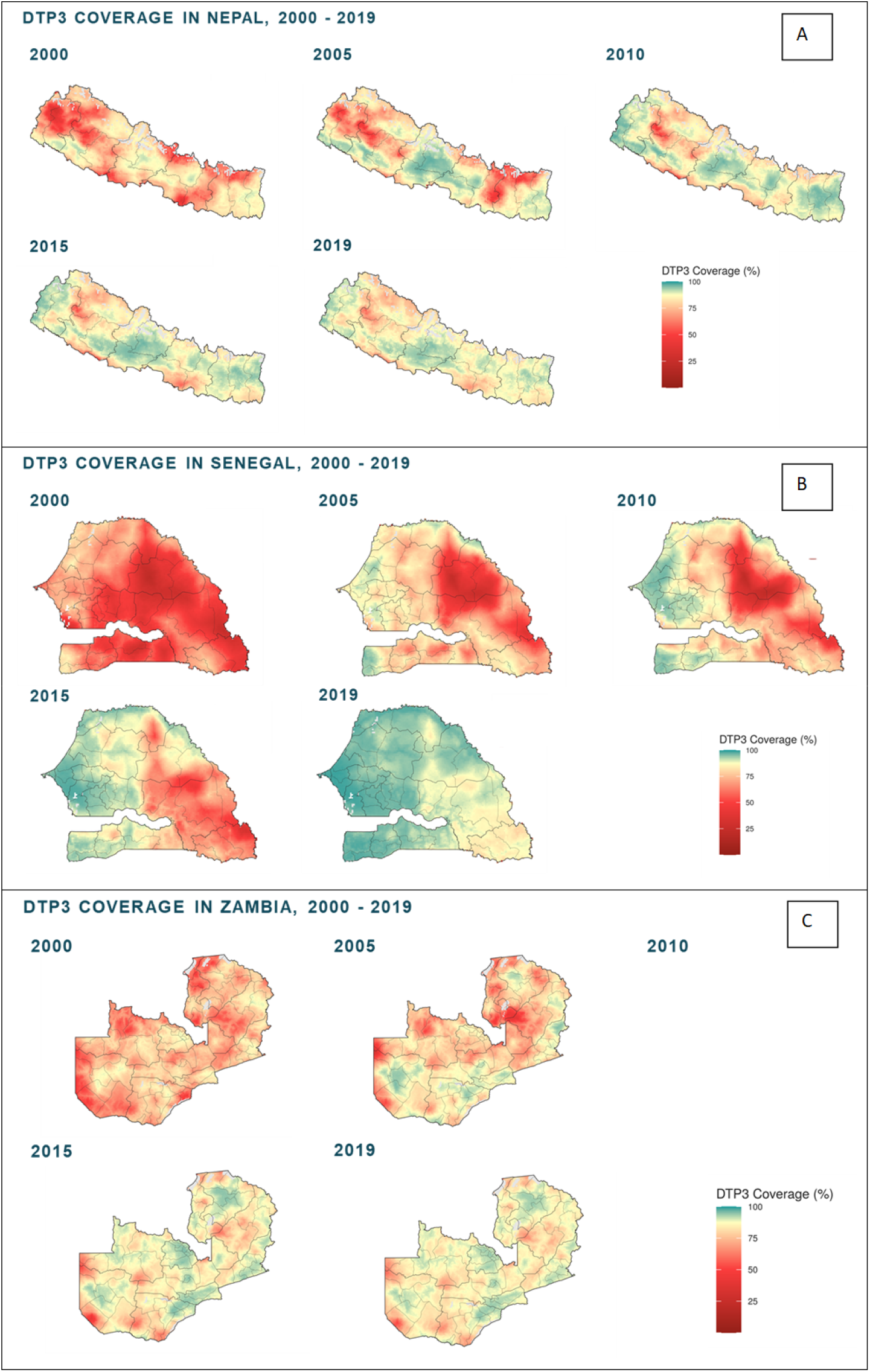
Historical patterns of sub-national DTP3 vaccine coverage in the three identified Exemplar countries: Nepal (Panel A), Senegal (Panel B), and Zambia (Panel C)

### In-Country Stakeholder Identification

Alongside our network of in-country and regional collaborators and networks, we identify a comprehensive list of key stakeholders to include in data collection. We aim to identify both individuals who were in the related positions at the time of data collection, and those who previously held such positions to assess how programmatic changes were implemented and adapted over time. The generalized list of positions is documented in Table 2; due to local context and health system structure, specific positions may differ by country. Specific categories and titles, and the number of related data collection activities, will be presented alongside country-specific analyses.

**Table 2.** General summary of key informant and focus group participants by roles within the vaccine system

| Method | Participant Position |
| --- | --- |
| Key Informant Interviews | Minister of Health, or other high-ranking officials (National) |
|  | Ministry of Education liaison (National) |
|  | Ministry of Finance liaison (National) |
|  | Partner organization officials (WHO, UNICEF, CDC, etc.) |
|  | Provincial/Regional health officers |
|  | District health officers |
|  | Health facility supervisors and nurses |
|  | Health unit workers (vaccinators, cold chain officers, etc.) |
|  | Community-based workers and volunteers |
|  | Community and religious leaders |
| Focus Group Discussions | Community health workers and volunteers |
|  | Mothers |
|  | Fathers |
|  | Grandmothers |

### External Advisory Group

We formed a Technical Advisory Group (TAG) consisting of experts in global health, vaccination delivery, vaccine confidence, and LIC and LMIC health systems to facilitate interpretation and dissemination of findings. The engaged stakeholder groups include WHO, UNICEF, CDC, and Gavi. Engagement of the TAG is an ongoing process, with meetings convened for discussion at key decision points - including, but not limited to, input on final country selection, review of preliminary findings, review of context around key findings, and the current development of plans for dissemination

### Conceptual Frameworks

This project uses several frameworks, which guided the development of tools and areas of inquiry. These overarching frameworks were taken from literature on vaccine delivery and implementation science. Implementation science is a growing field with the focus on applying evidence-based research findings into routine practice. Additional cross-cutting analyses utilize discipline-specific frameworks based on and extrapolated from the existing literature. The primary outputs of this study are country-level case studies, with additional cross-topic synthesis as possible.

#### Vaccine Delivery Framework

Our conceptual model organizes the complex interplay of barriers and factors impacting global childhood vaccine coverage, based on the work of Phillips et al.^[9]^ and LaFond et al.,^[10]^ and a broader review of the vaccine confidence and coverage literature (Figure 4). Specific input was provided by our multi-disciplinary team of public health, behavioral science, implementation science, political science, public policy, and systems science and engineering researchers. This novel framework serves as a guiding summary of the key issues for consideration in each country. The research is driven by the findings from each country (see Research Activities below), with no pre-conceptions regarding specific practices or interventions. An initial scoping visit for each exemplar country was used to gather preliminary feedback about the immunization program, historical challenges and interventions, and key stakeholders’ initial impressions about reasons for success. These findings were then compared to the overall framework in Figure 4 to identify specific areas in which additional focus was needed during the main research activities.

**Figure 4.**
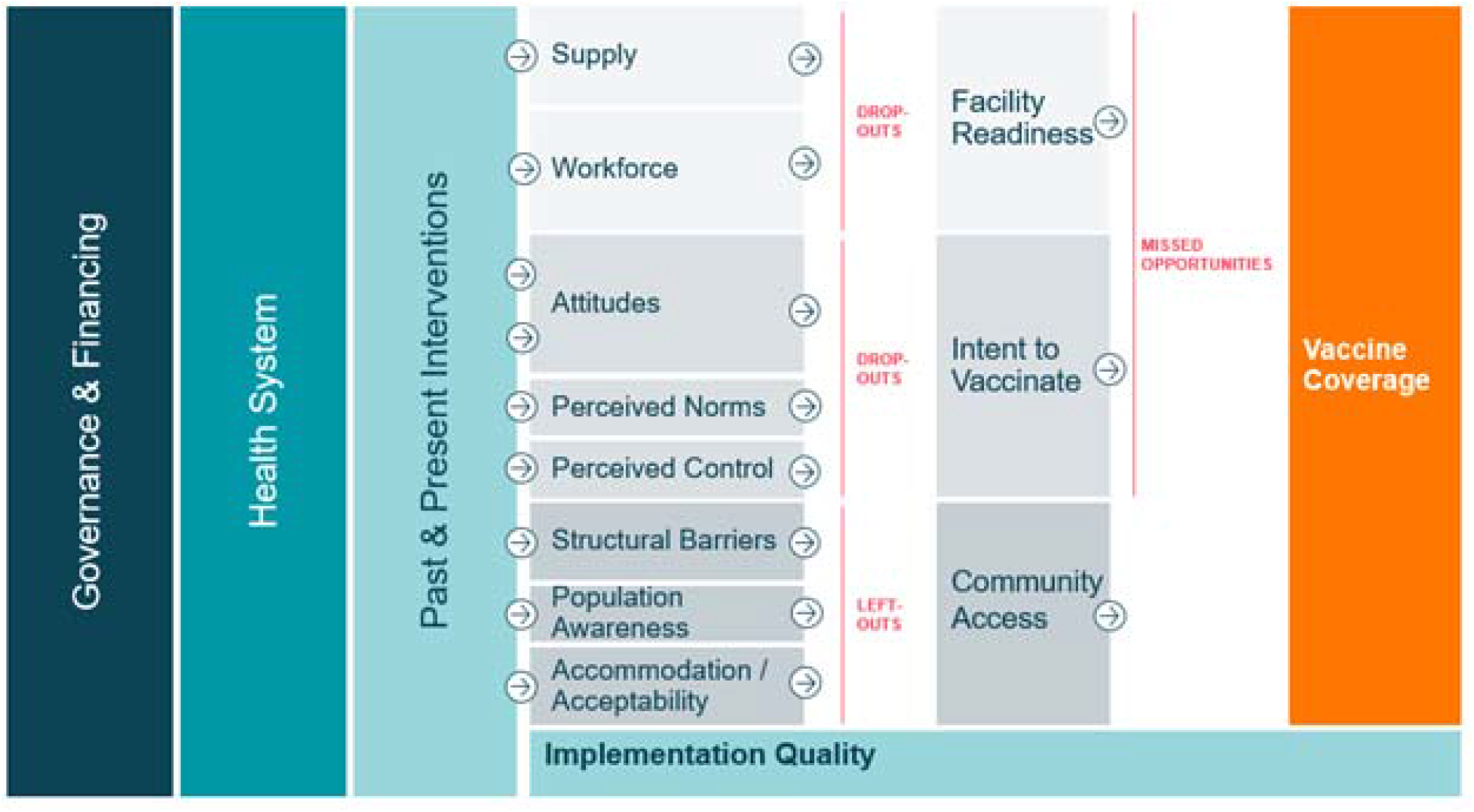
Conceptual framework of drivers of vaccine delivery, derived from scoping visits and Phillips, et al.^[9].^ *Note: Drop-outs refer to children who started their immunization series, but never completed. Left-outs refer to children who did not have access to immunization services. Missed opportunities refer to children with access to services who did not receive vaccinations when recommended.

#### Towards Developing Actionable Recommendations

The goal of this project is to provide evidence-based, actionable recommendations to country and global stakeholders, with a focus on new insights to exemplary performance of vaccine delivery. Our initial scoping visits identified key historical barriers and interventions in each country; the focus of this research is understanding the “how” and “why” related to the adoption of each of these interventions or activities. Interventions may have been developed by stakeholders within each country (i.e., endogenous innovation) or may be adaptations of higher-level guidance, such as local implementation of WHO guidance (i.e., exogenous adaptation). For each intervention or program – defined here as a solution developed and delivered by the country stakeholders (“what”) - there is an iterative process between identifying the problem to be addressed (“why”) and developing mechanisms for change, in other words “how” the change could come about (Figure 5). Understanding the interplay between “how,” “why,” and “what” can help identify actionable recommendations that may be useful for countries to consider when evaluating improvement in their vaccination systems.

**Figure 5.**
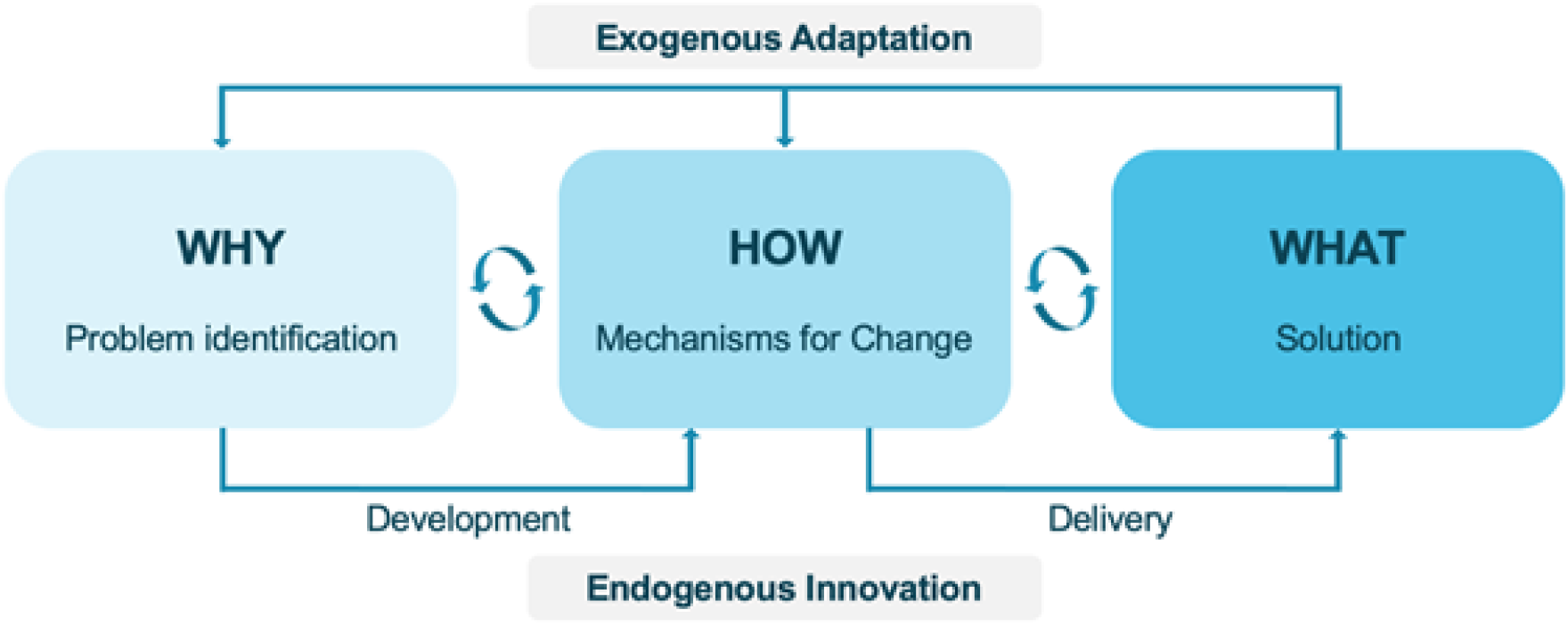
Mapping the “how” and “why” behind an intervention.

#### Implementation Science Frameworks

A combination of two implementation science frameworks was applied to develop tools for data collection. Application of these frameworks directed our inquiry towards key domains of the historical decision-making and implementation process.

##### Consolidated Framework for Implementation Research (CFIR)

CFIR is a framework of five interrelated domains (intervention, outer setting, inner setting, individual characteristics, and process of implementation) which influence the effectiveness of intervention implementation, and promote hypotheses of “what works where and why across multiple contexts.”^[16]^ We identified constructs within CFIR for focus within our tool development - including motivation, decision-making processes, mechanism for change, and the process and environment of development and delivery - in addition to inquires of events and policies most relevant to the success of Exemplar countries. The CFIR framework guides our examination of “what they did,” “why they did it,” and “how they did it,” at national, regional, district, and local levels in order to understand diverse contexts and perspectives within each of the exemplar countries. This allows us to systematically organize our findings, and better interpret the similarities and differences both across and between exemplar countries.

##### Context and Implementation of Complex Interventions (CICI)

The CICI framework was applied in addition to CFIR to address contextual factors and the inter-dimensionality missing from the CFIR framework; both framed our initial thinking about the vaccine delivery system. ^[17]^ Both CFIR and CICI frameworks guided the development of an iterative data collection tool that could be applied consistently across diverse contexts and settings.

### Research Activities

#### Tool Development

Qualitative data collection was guided by semi-structured key informant interview (KII) guides for use with health officials, external stakeholders, and community leaders, and focus group discussion (FGD) guides for use with fathers, mothers, grandmothers, and community health workers. These instruments explore the following CFIR and CICI domains: intervention characteristics, outer setting, inner setting, characteristics of individuals, process, and context.^[16]^ Qualitative data collection was intended to limit the time burden for KII or FGD participants to no longer than one hour, although some data collection took longer - up to two hours or more - based on the richness of the discussion. An initial KII guide was developed for scoping visits and was revised post visit to ensure data was captured within the domains of interest raised in those KIIs. Our overarching goal was to gather information from participants about “how” and “why” interventions were developed, adapted, and implemented, and how they led to an increase in vaccination coverage. The guides were developed by the research team and refined through iterative review after completion of data collection in each country.

#### Scoping Visits

Prior to beginning both in-depth data collection and review of relevant literature, we conducted a two-week scoping visit in each country to (a) meet with and select in-country partners; (b) discuss key factors of change for further exploration (e.g., identify the “what” items for exploration of “how” and “why”); and (c) prepare for in-depth country research activities (e.g., establish local partnerships, start ethical reviews, research activity logistics).

#### Research Visits and Qualitative Data Collection

We conducted ten-day training workshops with our local research partners prior to the start of data collection in each country. In addition to training on study materials and methodology, we reviewed the materials alongside our in-country research partners to aid in any translation and adjust content for country context.

We conducted both KIIs and FDGs, as appropriate, with data collection occurring at the national level, sub-national levels, and community stakeholders at sub-national levels (Table 2). KIIs and FGDs took place in offices, clinics, and community centers. All activities took place in a location deemed private, safe, and comfortable by the participants. Qualitative data collection activities were conducted in person with trained facilitators and note-takers, when possible. Conditions for in-person research relative to the COVID-19 pandemic necessitated adjustments to maximize the quality of data collection and participant and researcher safety.

FGDs consisted of 6-8 participants. FGDs were held in the communities, organized by type of participant (e.g., fathers will be in one group), and consisted of groups of fathers, mothers, grandmothers, and community health workers. Partner organizations or community health workers identified the FGD participants.

#### Qualitative Data Analysis and Management

With permission from KII and FGD participants, interviews were recorded to ensure capture of all information. Recordings were transcribed verbatim from the local language by local research assistants and translated to English manually, or translated using Google Translate (for French), with verification by a fluent bilingual speaker. All documents with transcriptions were only accessible to researchers named on the IRB. All transcribed documents required a code to access. All research files, recordings, and transcriptions in-country were saved on password-protected computers. Recordings were removed from recorders at the end of every day, deleted once uploaded onto password-protected computers and saved to HIPAA-compliant storage in folders only accessible to the study team. All recordings have been removed from computers and servers following transcription and verification of accuracy. Interviewees’ names and contacts were de-identified, and all information will be used without mentioning their names. Documents that may link participants to their identifier code will be stored in separate locations.

Data were coded using MAXQDA 20 (Berlin, Germany) and analyzed thematically by specific aim, research question, and framework-specific construct(s). The initial analysis for each country consisted of a case study, specific to that country, identifying the key drivers of improvements in vaccine coverage. This broad case study served as a starting point for more detailed topic-specific analyses and manuscripts. For key factors identified in multiple countries, a cross-country synthesis will be conducted to identify similarities and differences in implementation across study countries.

#### Quantitative Data Collection

Quantitative data was gathered through freely obtained information on Ministry websites or data given from Ministry or other partners, such as the WHO, UNICEF, and CDC. This quantitative analysis investigates vaccine coverage through a review of the health spending and economic growth trends from LICs and LMICs. Selected exemplar countries are compared to this grouping to determine what factors made exemplar countries stand apart from their peers. Analysis will use cross-country and multi-year mixed-effects regression models to statistically test financial, economic, development, demographic, and other country-level indicators. A key component of this research will be to identify factors that may have been associated with improvements in vaccine coverage that are not commonly used as indicators of immunization. This can include general health systems strengthening and improvements in funding for public health, as well as improvements in maternal and child health that may have driven support for immunization services.^[18]^

#### Patient and Public Involvement

We consulted with a technical advisory group, but did not directly solicit patient or public involvement in the development of this research project.

## Ethics and dissemination

### Ethics

The study was approved by the Emory University Institutional Review Board (IRB); the Nepal Health Research Council in Nepal; the University of Zambia Biomedical Research Ethics Committee and the National Health Research Authority in Zambia; and the Comité de National d’Ethique pour la Recherche en Santé (CNERS; National Ethical Committee for Health Research) in Senegal. Participation in KII or FGD was voluntary, and interviewees were asked to provide informed consent.

### Dissemination

In addition to country-specific manuscripts describing our learnings, we will generate recommendations for national-level immunization programs based on the findings from this project. Specific reporting structures are listed below.

1. **Country-level reports and case studies**. The investigators will produce country-level findings, with feedback from country-level stakeholders. Country-level case studies will provide the basis for peer-reviewed manuscripts and broad dissemination on the Global Exemplars web platform.
2. **Domain-level analysis**. We will analyze each domain of interest identified from country case studies; these domains will be explored across exemplar countries. Current domains of interest for this synthesis include: targeted disease control activities, roles of community health workers and volunteers, health spending across LICs and LMICs, and intent and demand for vaccines. Findings will be disseminated among key national and global stakeholders and will be submitted for peer-review publication and for dissemination of the Gates Ventures web platform as cross-cutting synthesis.
3. **Tool and protocol development**. All individual frameworks and tools used by the research teams to inform research from their individual disciplines will be publicly available.
4. **Knowledge translation and implementation outreach**. Regional technical advisory meetings, webinars, policy fora, academic conferences, the exemplars platform, and global partner meetings will be leveraged to disseminate findings. Additionally, findings will be translated into recommendations of replicable solutions for non-exemplar countries and areas for potential intervention investment for global immunization actors and policymakers. Documents might include policy briefs and infographics.
5. **Exemplars in Global Health website**. Exemplars.health is the platform documenting the work of the Exemplars in Global Health Project by Gates Ventures and will include narratives based on the research not just from the Vaccine Delivery project described here, but all other Exemplars in Global Health Projects.^[12]^ The research team is working collaboratively with Gates Ventures to iteratively translate the research findings to the platform for public consumption.

## Conclusions and Limitations

The Exemplars in Vaccine Delivery Project offers an opportunity to evaluate the critical factors in childhood vaccine delivery in LICs and LMICs. The in-depth qualitative data collection and analysis will provide a deeper understanding of this issue based on the experiences and perspectives of key leaders in the three exemplar countries. Quantitative findings and existing literature will be used to triangulate findings. Our multi-disciplinary team brings experience in the fields of vaccine hesitancy, vaccine program delivery, behavioral science, implementation science, public policy, political science, systems engineering. With a focus on changes over the previous two decades that may have spurred catalytic growth in vaccine coverage, these findings will present a unique opportunity to identify not just areas for improvement in global vaccine delivery, but the most appropriate methods to consider during implementation of these solutions. Longstanding efforts in health system strengthening offer a framework to build on, and the actionable recommendations that will arise from this project present a novel means to support the health of and protection from infectious diseases for children around the globe.

## Data Availability

Data available upon request.

## Acknowledgements

Our technical advisory committee provided important consultation and feedback on this project: Agnes Binagwaho (University of Global Health Equity), Laura Craw (Gavi), Carolina Danavaro (WHO), Anuranda Gupta (Gavi), Heidi Larson (London School of Hygiene and Tropical Medicine), Kate O’Brien (WHO), Helen Rees (Wits Reproductive Health and HIV Institute), Lora Shimp (John Snow, Inc), Aaron Wallace (CDC). Chin-En (King) Ai and Allison Wray contributed to the initial phase of this project.

## Authors’ contribution

The study was conceived of by MCF, RAB, SO, WO; The protocol was developed by MCF, RAB, CE, KAH, AE, BD, Country selection was guided by KAH, MCF, and RAB; Country-level regional selection and tool adaptation was led by SD, WK, MS, KM, BD, KAH, AE, ZS; The first draft was written by RAB with editing by MCF, KAH, AE, KM, ZS, WK, SD, and MS. Quantitative methods developed by PK, DN, FZ. All authors provided input to facets of the overall study design and reviewed and approved the final manuscript.

